# Social contacts and transmission of COVID-19 in British Columbia, Canada

**DOI:** 10.1101/2021.09.23.21263649

**Authors:** Notice Ringa, Michael C. Otterstatter, Sarafa A. Iyaniwura, Mike A. Irvine, Prince Adu, Naveed Z. Janjua, Samara David, Michelle Spencer

## Abstract

**Purpose:** Close-contact rates are thought to be a significant driving force behind the dynamics of transmission for many infectious respiratory diseases. Efforts to control such infections typically focus on the practice of strict contact-avoidance measures. Yet, contact rates and their relation to transmission, and the impact of control measures, are seldom quantified. Here, we quantify the response of contact rates, transmission and new cases of COVID-19 to public health contact-restriction orders, and the associations among these three variables, in the Canadian province of British Columbia (BC) and within its two most densely populated regional health authorities: Fraser Health Authority (FHA) and Vancouver Coastal Health Authority (VCHA).

**Methods:** We obtained time series for self-reported close-contact rates from the BC Mix COVID-19 Survey, new reported cases of COVID-19 from the BC Center for Disease Control, and transmission rates based on dynamic model fits to reported cases. Our study period was from September 13, 2020 to February 19, 2021, during which three public health contact-restriction orders were introduced (October 26, November 7 and November 19, 2020). We used segmented linear regression to quantify impacts of public health orders, Pearson correlation to assess the instantaneous relation between contact rates and transmission, and vector autoregressive modeling to study the lagged relations among the three variables.

**Results:** Overall, declines in contact rates and transmission occurred concurrently with the announcement of public health orders, whereas declines in new cases showed a reporting delay of roughly two weeks. The impact of the first public health order (October 26, 2020) on contact rates and transmission was more pronounced than that of the other two health orders. Contact rates and transmission on the same day were strongly correlated (correlation coefficients = 0.64, 0.53 and 0.34 for BC, FHA, and VCHA, respectively). Moreover, contact rates were a significant time-series driver of COVID-19 and explained roughly 30% and 18% of the variation in new cases and transmission, respectively. Interestingly, increases in transmission and new cases were followed by reduced rates of contact: overall, average daily cases explained about 10% of the variation in provincial contact rates.

**Conclusion:** We show that close-contact rates were a significant driver of transmission of COVID-19 in British Columbia, Canada and that they varied in response to public health orders. Our results also suggest a possible feedback, by which contact rates respond to recent changes in reported cases. Our findings help to explain and validate the commonly assumed, but rarely measured, response of close contact rates to public health guidelines and their impact on the dynamics of infectious diseases.

## 1 Introduction and literature review

A wide variety of infectious respiratory diseases, including influenza, measles, pertussis, plague, tuberculosis and the new Coronavirus Disease 2019 (COVID-19), are transmitted largely through close-contact and spread based on the social contacts and mixing patterns of the host population [4, 5, 10]. High rates of effective contacts (interactions that allow pathogen transfer between individuals) are often associated with increased risk of infection. Effective contacts typically involve inhalation of infectious secretions from coughing, sneezing, laughing, singing or talking, but may also include touching contaminated body parts or surfaces followed by ingestion of the pathogen [4, 14]. Control strategies against such infections are based on contact avoidance measures, including isolation of those who are ill, use of personal protective equipment such as gloves and face masks, and physical distancing [27, 19]. In this study we examine the relation between self-reported social contact patterns, public health control measures, and the transmission of COVID-19 in the province of British Columbia (BC), Canada. BC is the westernmost province in Canada with a population of 5.2 million, of which about 62% live within the greater metropolitan area of the lower mainland which comprises the regional health authorities Fraser Health Authority (FHA) and Vancouver Coastal Health Authority (VCHA) [16].

COVID-19, a viral respiratory infectious disease caused by Severe Acute Respiratory Syndrome Coronavirus 2 (SARS-CoV-2) [2, 9, 11, 6], emerged in Wuhan, China in 2019 [45, 24] and has since spread worldwide to over 220 million people and led to over 4.5 million deaths [15]. The primary mode of transmission for COVID-19 is contact with and inhalation of infectious respiratory droplets, which may propel two meters or more before settling on environmental surfaces [6]. Symptoms of COVID-19 include fever or chills, cough, shortness of breath, fatigue, muscle or body aches, headache, loss of taste or smell, sore throat, which may result in mild to severe illness, and death [25]. Governments around the world have attempted to contain the spread of COVID-19 by restricting social gatherings, closing schools, suspending recreational activities and travel, and by other means of reducing contacts. Nevertheless, the impacts of public health measures on actual rates of close contact, and the relation between these contacts and transmission, are rarely quantified.

A small number of studies, including in [26, 33, 23, 28], have analyzed population patterns of social contacts, and their connection to the dynamics of close-contact infectious diseases. The studies found that disease incidence and effective reproduction number (average number of newly infected individuals per case) increase with contact rates. However, contact rates and their effects on the infection dynamics vary over time and with epidemiologically critical factors such as geographical location, sex, age, household size, occupation and other socio-economic factors. For instance, large households increase density, which promotes opportunities for infection and also potentially expose individuals to a wider distribution of age-groups such as in multi-generational households.

In our study, we explore and quantify associations between social contact patterns, public health orders, disease transmission, and reported cases of COVID-19, in BC. We make use of detailed contact survey data and estimate transmission using a model-based metric of the time-varying reproductive number, *R*_*t*_. We specifically consider data from autumn of 2020 onward, during which a series of regional and provincial public health orders were introduced to reduce the number of close contacts and curb transmission. We compare associations at the provincial level, with those from the two largest regional health authorities of BC: FHA and VCHA. We hypothesize that rates of contact (as measured by the average number of self-reported close-contacts made by an individual in a day) are predictive of the instantaneous *R*_*t*_ estimates and drive the spread of COVID-19 within a region.

## 2 Methods

We studied the association between contact rates and transmission of COVID-19 in BC from September 13, 2020 to February 19, 2021, a period in which three public health contact-restriction orders were introduced (October 26, November 7 and November 19). For each successive four-day period, we calculated (i) population rates of contact as the average number of self-reported close-contacts made by an individual in a day (average daily contacts); (ii) the average number of newly reported COVID-19 cases per day (average daily cases or new cases); and (iii) transmission rate of COVID-19 as the average daily value of our model-based estimate of *R*_*t*_. Our *R*_*t*_ indicator was derived by fitting the *covidseir* transmission model of [2], where *R*_*t*_ was computed using the Next-Generation matrix method [8, 29], to the reported case data. We analyzed the time series of these three variables and used linear segmented regression to quantify the change in trend associated with announcement of public health orders. We used Pearson correlation to assess the instantaneous relationship between average daily contacts and transmission (*R*_*t*_), and used vector autoregressive (VAR) models to quantify the lagged associations between average daily contacts and average daily cases, and *R*_*t*_. All analysis was performed using R version 3.6.3.

### 2.1 Data

The British Columbia COVID-19 population mixing patterns survey (BC-Mix) (http://www.bccdc.ca/our-research/projects/bc-mix-covid-19-survey) is an ongoing online survey launched by the British Columbia Centre for Disease Control (BCCDC) on September 04, 2020. These survey data are used to measure and assess contact patterns and activity levels in BC during the COVID-19 pandemic and to inform the timing of easing and re-imposing physical distancing measures. The survey comprises 94 items across six key domains, namely demographic information; COVID-19 testing and results, symptoms, and health behaviours; activities and behaviour in and outside of the home; internet and social media use; perceptions and attitudes around COVID-19; and COVID-19 vaccine acceptance. Survey participants are aged 18 years and above. A detailed description of the survey including its development, design, case definitions and other characteristics is described in [21]. Survey respondents record numbers of close contacts made in a single day, in answer to the question “ How many people did you have in-person contact with between 5 am yesterday and 5 am today?”. In the survey, in-person contact is defined as face-to-face two-way conversation with three or more words, or physical skin-to-skin contact such as a handshake, hug, kiss and contact sports. For the purposes of this study, we used weighted survey data including only the “baseline” responses (i.e., participants can complete the survey multiple times, but we included only the first completed survey for each individual). Weighting was used to correct for differences between the distribution of respondents (by age, sex, geography and ethnicity) and that of the BC population.

BC COVID-19 case data, which is available at [31], provided numbers of daily confirmed new cases of COVID-19 in BC, based on case report date and symptom onset date. COVID-19 case data are collected for BC residents by the regional health authorities during public health follow-up and reported to the BCCDC [18]. The BCCDC compiles the regional data into a provincial dataset [31], which includes the health authority and date reported to public health (or symptom onset date, when reported date is not available) for each case.

### 2.2 Contact restrictions

The first case of COVID-19 in BC was detected on January 28, 2020, after which rapid spread of the virus led to a number of public health contact-restriction orders [15, 31]. At various times, these orders included temporary reduction in the sizes of social gatherings, avoidance of non-essential travel, strict physical distancing, cancellation of sporting activities, shut-down of businesses, school closures, work-from-home arrangements, mandatory indoor masking and quarantine for travelers. These orders were associated with marked declines in transmission after the initial peak in spring of 2020 (the so-called ‘first wave’), with only 30-40 new cases reported daily in BC throughout the summer of 2020. Nevertheless, cases grew rapidly during autumn of 2020 (the ‘second wave’): during November alone, the number of newly reported cases quadrupled, from roughly 200 per day to 800 cases per day. On October 26, 2020, the province restricted social gatherings to household members plus their immediate six close-contacts; on November 07, 2020, additional restrictions limited social gatherings to household members only for the two most populous regional health authorities (FHA and VCHA); and on November 19, 2020, mandatory indoor masking was announced and social gatherings were limited to household members only for the entire province. These restrictions were followed by a steady decline in newly reported COVID-19 cases during December 2020.

### 2.3 Statistical analysis

We used linear segmented regression models to investigate the impact of public health orders on the time series for average daily contacts, new cases and *R*_*t*_. We used standard Pearson correlation (described in Appendix B) to assess if high contact rates and high transmission (*R*_*t*_) tended to occur at the same time. We compared these correlations between BC and the FHA and VCHA regional health authorities. Following this analysis, we conducted time series (VAR) modeling- to determine if transmission and new cases were affected over time by contact rates. We use *α* = 0.05 for all statistical tests in this paper.

#### 2.3.1 Segmented regression

Segmented (or piecewise) regression is a method of time series analysis in which the independent variable (time) is divided into a number of intervals or segments, which are connected via vertices called breakpoints or knots, and a separate line segment is fit into each interval [12, 37]. A model with *k* breakpoints divides a time series into *k* + 1 intervals. The general description of segmented regression models is given in Appendix A. In our analysis, regression breakpoints were located at the three dates on which public health orders were officially announced. Therefore, the equation of each of the regression lines in Figure 1 takes the form

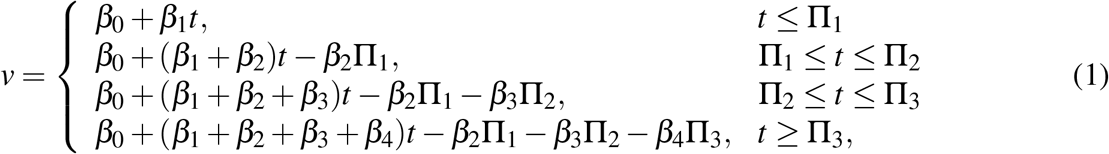

where *v* = *v*(*t*) is the regression-estimated value of a variable *v* at time *t*; Π_1_, Π_2_ and Π_3_ are the dates of announcement of the first, second and third public health order, respectively; *β*_0_, *β*_1_, *β*_2_, *β*_3_ and *β*_4_ are constants to be estimated based on time series data. According to Equation (1), for each regression line in Figure 1, the slope of the first, second, third and fourth segment is given by *β*_1_, *β*_1_ + *β*_2_, *β*_1_ + *β*_2_ + *β*_3_ and *β*_1_ + *β*_2_ + *β*_3_ + *β*_4_, respectively. Thus, *β*_2_ represents the change in the regression slope from the interval *t* ≤ Π_1_ to Π_1_ ≤ *t* ≤ Π_2_; *β*_3_ represents the change in the regression slope from the interval Π_1_ ≤ *t* ≤ Π_2_ to Π_2_ ≤ *t* ≤ Π_3_ and *β*_4_ represents the change in the regression slope from the interval Π_2_ ≤ *t* ≤ Π_3_ to *t* ≥ Π_3_. We fit linear segmented regression models to our time series and generated estimates (i.e. *β*_0_, *β*_1_, *β*_2_, *β*_3_ and *β*_4_) for each regression line using the *segmented* R-package (see Appendix A.1).

**Figure 1.**
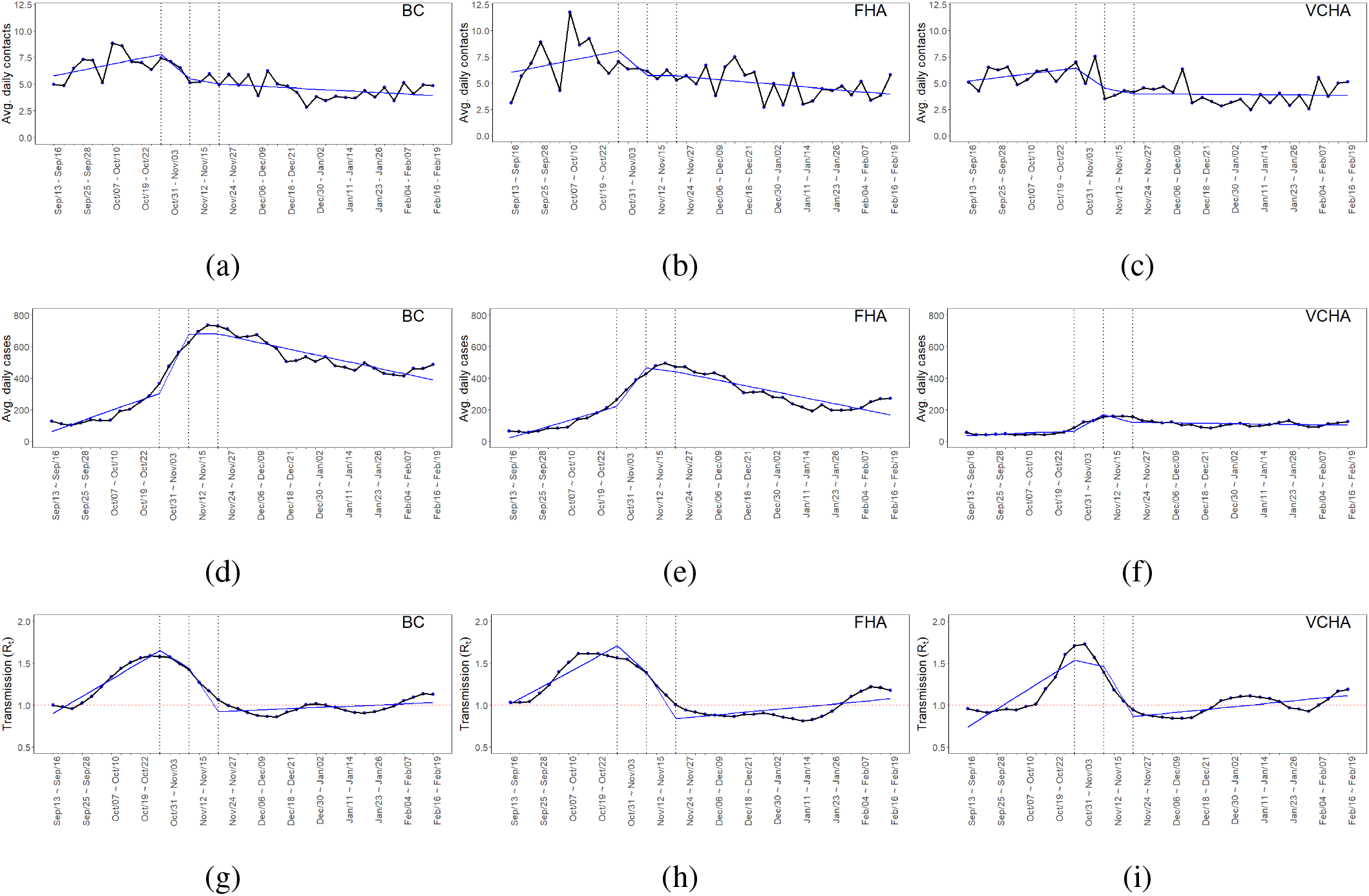
Time series of average daily contacts (contact rates), average daily cases (new cases) and transmission (*R*_*t*_) of COVID-19 in BC (1a, 1d and 1g), FHA (1b, 1e and 1h) and VCHA (1c, 1f and 1i). The vertical dotted lines in each graph indicate the dates of announcement of public health contact-restriction orders on October 26, 2020, November 07, 2020 and November 19, 2020.

### 2.3.2 Vector autoregressive models

Autoregressive (AR) models are a time-series analysis method that considers a variable in relation to its own past (lagged) values in order to determine significant associations over time and/or to predict future values [46]. Vector autoregressive (VAR) models are an extension of this approach and used to study relationships between two or more time-series variables, such that each variable is modeled in terms of its own past values as well as the past values of the other variables [46, 48, 42, 39]. In this paper, the time-series variables of interest are average daily contacts, average daily cases (new cases) of COVID-19 and average daily value of *R*_*t*_ (transmission). The general VAR model of *n* dependent variables with independent variable *t* and *l* lags, is given by

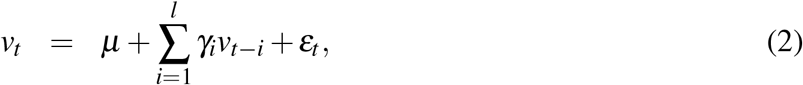

where *v*_*t*_ is an *n* × 1 vector representing time series variables at time *t*; *µ* is an *n* × 1 vector of constants (intercepts with the *v*_*t*_-axis) of time series; *γ*_*i*_ (*i* = 1, 2, …, *l*) is an *n* × *n* matrix of coefficients of lagged variables at time *t* − *i*; *ε*_*t*_ is an *n* × 1 vector of error terms (white noise) [46]. Estimation of a VAR model mainly involves approximation of the vector *µ* and coefficients *γ*_*i*_ that best describe relationships between different the time series, and determine the equation of each variable as a function of other variables in the system.

VAR models are used to provide estimates of stationary time series (stationary processes). A stationary process has no time-trends in the mean and variance of the data. Our time series shown in Figure 1 were detrended by differencing to achieve stationarity (confirmed by Augmented Dickey-Fuller (ADF) tests). Further details on differencing and ADF test can be found in Appendix C.1. Steps for fitting VAR models to time series data include selection of suitable lag lengths for model variables; estimation of model parameters and coefficients; testing of time series causality; and forecast error variance decomposition (FEVD) of variables. A detailed explanation of the VAR model fitting process can be found in Appendix C.2. Our VAR models for average daily contacts and new cases, and average daily contacts and transmission of COVID-19 in BC, FHA and VCHA, are derived in Appendix C.3.

Granger-causality is a useful method for capturing statistical relationships between VAR model variables and estimates the degree to which one time series predicts future values of another time series [46, 43, 44, 47]. The statistical test for Granger-causality compares the predictive ability of the time series model of interest with and without the putative causal variable and is therefore interpretable as evidence towards a causal relationship, but not as proof of a causal relationship. A variable *v* is said to Granger-cause a variable *w* if p-value, p, of the Granger-causality test is less than 0.05, otherwise, *v* does not Granger-cause *w*. FEVD is a statistical tool for measuring the amount of variation in a variable over time that is attributable to the variable’s own past values versus past values of other model variables [46]. Based on a fitted VAR model, FEVD plots illustrate variation over time prospectively, i.e., as a forecast. We present both FEVD and Granger-causality tests for our time series models in order to assess to what degree, and over what time frame, contact rates are driving changes in COVID-19 transmission and new cases.

## 3 Results

### 3.1 Effects of public health orders on average daily contacts, average daily cases and transmission

Provincially, rising contact rates and transmission (*R*_*t*_) reversed shortly after the first health order on October 26, 2020 (Figures 1a and 1g); for contacts, this declining trend lasted only until the second public health order (13 days later, on November 7), whereas for *R*_*t*_, the decline continued to at least the third order (25 days later, on November 19). Both contact rates and *R*_*t*_ were relatively stable after the third order until the end of our study period (February 19, 2021). As expected, the trend in new cases mirrored that of our transmission indicator but was shifted about two weeks later, corresponding to the delay between transmission to symptom onset followed by diagnosis, and case reporting (Figure 1d vs. Figure 1g). The same patterns were generally apparent in both of the regional health authorities we studied, although declines in contact rates and *R*_*t*_ appeared to start roughly one week before the first public health order in FHA, and roughly one week after the first order in VCHA (Figures 1b-1i). Simple comparison of overall contact rates and *R*_*t*_ before and after the introduction of public health orders indicated that in BC, FHA and VCHA, contact rates declined by 30.1%, 29.2% and 29.9%, while *R*_*t*_ declined by 17.9%, 25.0% and 5.40%, respectively, following the first public health order onwards.

To quantify impacts of each public health order, we fit linear segmented regression models to all time series in Figure 1, and plotted the corresponding regression lines with three knots situated at the three dates of introduction of public health orders. The equation of each regression line and model estimates are presented in Appendix A.1. In BC, FHA and VCHA, the slope of the contact rate regression line was positive before the first public health order, turned substantially negative thereafter and slightly increased, but remained negative through all other health orders (Table 1). The changes in contact rate slope after the first public health order (i.e. Π_1_ ≤ *t* ≤ Π_2_) were statistically significant in the province and in VCHA (p < 0.05), but not in FHA. Provincially and in the two regional health authorities, the changes in contact rate slope following the second and the third health orders (i.e. Π_2_ ≤ *t* ≤ Π_3_ and *t* ≥ Π_3_) were not statistically significant (p > 0.05). Provincially and in the two regional health authorities, the slope for transmission (*R*_*t*_) was positive before the first public health order, turned negative after this order, decreased further following the second public health order, and stabilized after the third health order (Table 1). Changes in transmission slope following all public health orders were statistically significant (p < 0.05), except after the second health order in FHA.

**Table 1:** Slopes of regression lines of average daily contacts and transmission in the province and in FHA and VCHA, within the four time intervals separated by the three dates (Π_1_, Π_2_ and Π_3_) of announcement of public health orders, based on associated model estimates *β*_1_, *β*_2_, *β*_3_ and *β*_4_ presented in Tables 3 and 5.

| | $t \leq \Pi_1$ | $\Pi_1 \leq t \leq \Pi_2$ | $\Pi_2 \leq t \leq \Pi_3$ | $t \geq \Pi_3$ |
| --- | --- | --- | --- | --- |
| Slope of BC average daily contacts | 0.184** | -0.768*** | -0.159 | -0.048 |
| Slope of FHA average daily contacts | 0.185 | -0.779* | -0.013 | -0.079 |
| Slope of VCHA average daily contacts | 0.111 | -0.634** | -0.182 | -0.007 |
| Slope of BC transmission | 0.068*** | -0.071*** | -0.173*** | 0.005*** |
| Slope of FHA transmission | 0.063*** | -0.105*** | -0.184 | 0.011*** |
| Slope of VCHA transmission | 0.072*** | -0.025*** | -0.199*** | 0.011*** |
| *Note:* *p<0.1; **p<0.05; ***p<0.01 |  |  |  |  |

### 3.2 Pearson correlation of average daily contacts and transmission

We used correlation analysis to identify if contact rates and transmission were concurrently related, i.e., if high contact rates and high transmission tended to occur at the same time. Provincially, and in both regional health authorities, transmission (average daily *R*_*t*_) was significantly positively correlated with average daily contacts (*r*^*BC*^ = 0.64, p < 0.001); *r*^*FHA*^ = 0.53, p < 0.001; *r*^*VCHA*^ = 0.34, p = 0.033). Based on these values, the magnitude of the correlation was about 50% stronger in FHA compared to VCHA (*r*^*FHA*^ ≈ 1.56 ×*r*^*VCHA*^).

### 3.3 VAR models of average daily contacts and average daily cases, and average daily contacts and transmission

Stationary processes for time series in Figure 1 are presented in Figure 3. The notations *BC*_*contacts*_*t*_, *BC*_*cases*_*t*_ and *BC*_*transmission*_*t*_ represent stationary processes of average daily contacts, average daily cases and transmission, respectively, in BC. The corresponding notations for FHA and VCHA are similarly defined. The selection of suitable lag lengths and derivation of equations of our VAR models of average daily contacts and average daily cases, and average daily contacts and transmission, are presented in Appendix C.3. Model estimates for all our VAR models are presented in Appendix C.4.

FEVD indicated that variation in new cases and transmission of COVID-19 were in part attributable to past values of average daily contacts, whereas variation in average daily contacts was explained largley by its own past values (Figure 2). Each panel shows the proportion of variation in cases, contacts or transmission that is explained by that variable’s own past values versus the past values of other variables. These panels illustrate changes over time as a forecast based on our VAR model, with each bar representing a period of four days.

**Figure 2.**
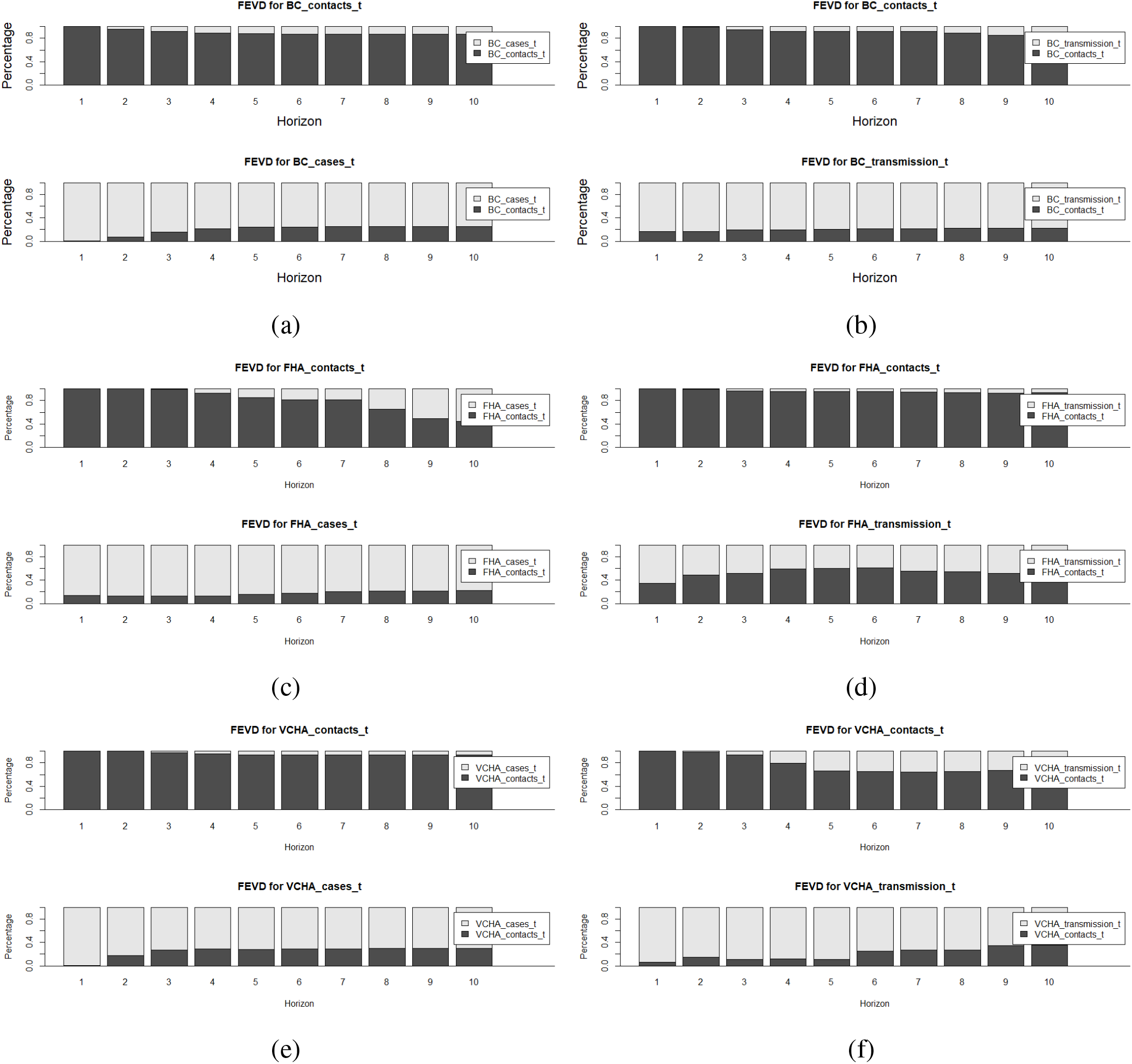
Forecast error variance decomposition results for VAR models of average daily contacts and cases, and average daily contacts and transmission in BC (2a and 2b), FHA (2c and 2d), and VCHA (2e and 2f).

Provincially, in the first eight days, past average daily contacts barely contributed towards variation in average daily cases, but from the third week onwards, about 30% of the variation in average daily cases was explained by previous average daily contacts (Figure 2a). Our time series analysis suggests that provincially, at all time periods, about 18% of the variation in COVID-19 transmission was explained by average daily contacts (Figure 2b). In FHA, at all time periods, previous average daily contacts contributed about 18% of the variation in average daily cases (Figure 2c) and an average of 50% of the variation in transmission (Figure 2d). In VCHA, from the end of the first week onwards, about 30% of the variation in average daily cases was explained by average daily contacts, whereas the contact rates explained up to 35% of the variation in transmission in the second month (Figures 2e and 2f).

Interestingly, FEVD also showed that, at relatively long time horizons, some variation in average daily contacts was explained by previous average daily cases and transmission of COVID-19. Provincially, from the third week, previous values of average daily cases and transmission explained up to 15% and 18%, respectively, of the variation in average daily contacts (Figures 2a and 2b). In FHA, past average daily cases contributed gradually to the variation in average daily contacts from the middle of the third week with up to 55% of the variation in the contact rates attributable to previous average daily cases in the sixth week (Figure 2c). From the beginning of the second month, previous transmission rates contributed less than 3% to the variation in average daily contacts in FHA (Figure 2d). In VCHA, previous average daily cases explained about 3% of the variation in average daily contacts from the second week, while transmission explained up to to 35% of variation in average daily contacts from the third week (Figure 2e and 2f).

Granger causality testing confirmed that provincially, previous values of daily contacts were a significant time series driver of average daily cases (p = 0.006), whereas the relatively small amount of variation in average daily contacts that was explained by previous case counts was only marginally significant (p = 0.049) (see Table 2). Granger causality testing also showed that in FHA, past average daily cases were a significant driver of average daily contacts (p = 0.001), but previous average daily contacts did not significantly drive time series for average daily cases (p > 0.05). In VCHA, average daily contacts were a significant time series driver of average daily cases (p = 0.011), whereas the reverse is false. Average daily contacts and transmission were not significant drivers of each other in the province and in FHA, but the two variables significantly drove time series of each other in VCHA.

**Table 2:** Granger causality test results for average daily contacts and average daily cases, and average daily contacts and transmission, in BC and two health regions, FHA and VCHA.

|  |  |
| --- | --- |
| $BC_{contacts\_t}$ G-causes $BC_{cases\_t}$ ($p = 0.006$) | $BC_{contacts\_t}$ does not G-cause $BC_{transmission\_t}$ ($p = 0.945$) |
| $BC_{cases\_t}$ G-causes $BC_{contacts\_t}$ ($p = 0.049$) | $BC_{transmission\_t}$ does not G-cause $BC_{contacts\_t}$ ($p = 0.544$) |
| $FHA_{contacts\_t}$ does not G-cause $FHA_{cases\_t}$ ($p = 0.519$) | $FHA_{contacts\_t}$ does not G-cause $FHA_{transmission\_t}$ ($p = 0.574$) |
| $FHA_{cases\_t}$ G-causes $FHA_{contacts\_t}$ ($p = 0.001$) | $FHA_{transmission\_t}$ does not G-cause $FHA_{contacts\_t}$ ($p = 0.582$) |
| $VCHA_{contacts\_t}$ G-causes $VCHA_{cases\_t}$ ($p = 0.011$) | $VCHA_{contacts\_t}$ G-causes $VCHA_{transmission\_t}$ ($p = 0.017$) |
| $VCHA_{cases\_t}$ does not G-cause $VCHA_{contacts\_t}$ ($p = 0.537$) | $VCHA_{transmission\_t}$ G-causes $VCHA_{contacts\_t}$ ($p = 0.023$) |

## 4 Discussion and conclusion

The primary approach to prevent the spread of many infectious diseases transmissible through close person-to-person contact is reduction or avoidance of such contacts altogether. Yet, few studies have quantified the impact that such contact-restrictions have on rates of ‘effective’ contact (those actually involved in transmission) and on transmission itself. In our study, we explored time series relationships between close contact patterns and the dynamics of the ongoing COVID-19 pandemic in British Columbia, Canada and in its two most populous regional health authorities, FHA and VCHA, from mid-September, 2020 to mid-February, 2021. During this period, three public health contact-restriction measures were introduced (on October 26, November 7 and November 19) to control rising numbers of cases. We used data from the BC Mix Survey, which specifically captures rates of close contacts that are likely to underlie transmission. We analyzed contact rates in relation to the timing of contact-restriction measures and assesed their impact on COVID-19 transmission (average daily number of new infections generated per case, *R*_*t*_) and reported new cases.

We found that in BC, FHA and VCHA, all the three public health orders reduced contact rates, transmission and subsequent new cases of COVID-19. Overall, declines in contact rates and transmission occurred concurrently with the announcement of public health orders, whereas declines in newly reported cases were, as expected due to reporting delays, lagged by roughly two weeks. The decline we observed in contact rates in FHA about one week prior to the public health orders could have resulted from public anticipation and early news reporting of the upcoming restriction orders and/or reports of rising numbers of new cases of COVID-19, particularly in the FHA region. In all regions, transmission curves mirrored, and were significantly correlated with those of contact rates, suggesting that these self-reported rates of close contact were directly related to spread of COVID-19. Contact rates declined by roughly 30% overall after the first public health order. Transmission similarly declined in response to these orders, although this effect varied by region (*R*_*t*_ reduced by 17.9%, 25.0% and 5.40% in BC, FHA and VCHA, respectively). The strong positive correlation between contact rates and transmission correlation coefficients: *r*^*BC*^ = 0.64, *r*^*FHA*^ = 0.53, and *r*^*VCHA*^ = 0.34, (p < 0.05 in all cases) suggests a contemporaneous relation between these two quantities, i.e. changes in contacts were directly associated with changes in transmission. Through time series analysis, we show that average daily contacts significantly predict average daily cases and transmission of COVID-19, and explained roughly 30% and about 18% of the variation in subsequent new cases and transmission, respectively, at the provincial level. Interestingly, we also found evidence of a possible feedback, with increasing transmission and numbers of new cases tending to result in decreased subsequent rates of contact: overall, average daily cases explained less than 15% of the variation in average daily contacts in the province. The interdependence of previous contact rates, new cases and transmission of COVID-19 varied by region.

A few studies have quantified variation in transmission or cases of an infectious disease as a function of contact rates. For instance, in [33], the authors analyzed United Kingdom contact survey data during periods before and after the March 2020 lockdown due to the COVID-19 pandemic, and found that a model-derived effective reproduction number declined by 75% as a response to a 74% reduction in average daily contacts. In [26], the authors studied contact survey data from Belgium during different stages of intervention against COVID-19 and found that an 80% decline in the average number of contacts during the first lockdown period resulted in a decline of the effective reproduction number to below one, resulting in fewer reported new cases. In [40], the authors studied United Kingdom population mixing patterns during the 2009 H1N1 virus influenza epidemic and found that a 40% reduction in contacts among school children during school holidays resulted in about 35% decline in the reproduction number of influenza. These studies confirm a relation between self-reported contact rates and infectious disease transmission, but also show variation that may be due to epidemiological factors such as difference in the transmission environment (e.g. use of personal protective equipment) and the types of contacts being measured.

The observed feedback mechanism in which contacts rates can decrease as a result of increasing transmission and new cases, has also been documented in a number of previous studies. For instance, during the 2014 Ebola outbreak in Sierra Leone, self-reported prevention practices such as avoidance of contacts with corpses, were found to have increased with rising disease prevalence [7]. During the early stages of the COVID-19 pandemic, the practice of cautious social contacts by the Singaporean population, increased with rising rates of infection due to behavioural drivers such as fear and perceived risk of infection [13]. Similarly, the decline of close contacts in Hong Kong during the first quarter of 2020 is thought to have resulted from increasing messaging and spread of information about the prevalence of COVID-19 [49]. Thus, wide-spread public awareness of increasing numbers of new cases, through public health and various information media, may help to explain population reductions in contact rates.

In our study, we found that contact patterns and the related dynamics of COVID-19 varied with the geographies considered. A number of previous studies have also identified variation in contact rates by geography, and by factors that themselves vary geographically. In [23], the authors analyzed and compared social contact survey data for eight European countries in 2005 and 2006, and found that contact rates varied by geographical location, but also by sex, age and household size. In [41], the authors reviewed contact survey data across several countries from varying economic brackets and found that, in general, high contact rates were associated with densely populated settings and large household sizes, which characterized most low to middle-income countries. This is consistent with the general expectation that close-contact infectious diseases are more likely to impact densely populated regions and settings with large household sizes. Geographic variation in our results, particularly the higher contact rates, transmission and numbers of new cases in FHA compared to VCHA, may reflect the generally higher population density and larger household sizes in FHA [16, 52].

Our analysis has several important limitations. We relied on case surveillance data to determine the number of new cases and the transmission indicator of COVID-19 over time. This means we did not account for asymptomatic infection, which may be a strong driver of COVID-19 transmission, and could have impacted the conclusions of our study. Relying on case surveillance data may also underestimate the actual number of new cases in settings where symptomatic individuals did not seek testing or where testing capacity is constrained by inaccessibility or shortage of supply of resources. Three regional health authorities were not included in the assessment of regional associations of contact rates to COVID-19 dynamics - the Northern, Interior and Vancouver Island Health Authorities. These health authorities have relatively smaller population sizes, are sparsely populated with many rural communities and have reported smaller numbers of COVID-19 cases, which may have also been underestimated due to limited testing in some remote communities [16, 31]. There were not enough self-reported contact rate data points to explore relations between close contacts and COVID-19 dynamics for these regional health authorities during the period that we studied. As a result, this study may underrepresent rural populations in BC. Limitations of the survey of self-reported contact rates that may affect our analysis, are provided in [Prince_paper]. For instance, some population groups including the economically marginalized, the unhoused and those in immigration detention or imprisoned, are underrepresented in the survey possbibly due to lack of access to social media, computer or eletronic devices and the internet, which were used to advertise and complete the survey. Because the epidemiology of COVID-19 is an emerging area of study, there may be other important unknown factors that can bring about additional limitations for our analysis.

To our knowledge, our study provides the first quantitative approach to measuring the associations between self-reported close contact rates, to public health contact-restriction orders and transmission dynamics of the COVID-19 in Canada. The observed positive correlation between contact rates and transmission of COVID-19 as well as the strong capability of contact rates to drive the spread of COVID-19 are likely to prevail, although with varying magnitudes, in BC, other Canadian or international health jurisdictions, and in the context of future similar infections that transmit by the respiratory or close-contact route. The findings advance the understanding of the quantitative value of contact rates, which can used to inform infectious disease control strategies.

## Data Availability

All data referred to in the manuscript are available at British Columbia Centre for Disease Control (www.bccdc.ca).

## Appendix

### A Segmented regression

The general linear segmented regression model of a dependent variable *v* and independent variable *t* with *k* knots Π_1_, …, Π_*k*_, is given by

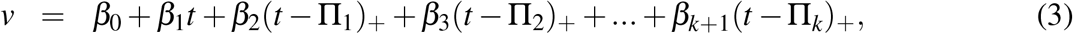

where *β*_0_, *β*_1_, *β*_2_,…,*β*_*k*+1_ are constants to be determined, and the terms (*u*)_+_ have the value *u* if *u* is positive, and 0 otherwise [37, 38]. Equation (3) can also be written as

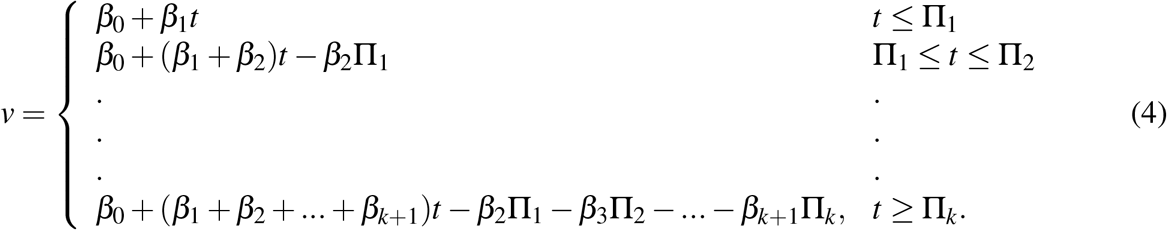

Thus, the *v*-intercept and slope of the first segment are *β* _0_ and *β*_1_, respectively; the intercept and slope of the second segment are *β*_0_ − *β*_2_Π_1_ and *β*_1_ + *β*_2_, respectively; and the intercept and slope of the *k* + 1^*th*^ segment are *β*_0_ − *β*_2_Π_1_ − *β*_3_Π_2_ − … − *β*_*k*+1_Π_*k*_ and *β*_1_ + *β*_2_ + … + *β*_*k*+1_, respectively.

#### A.1 Estimates for linear segmented regression lines in Figure 1

This section presents tables of estimates (i.e. *β*_0_, *β*_1_,…, *β*_4_) for linear segmented regression lines in Figure 1, where the breakpoints, Π_1_, Π_2_ and Π_3_, are located at the dates of the three BC public health orders on October 26, 2020, November 07, 2020 and November 19, 2020. The number of ‘*’s besides each estimate increases with the statistical significance of the estimate, and the values inside brackets () are standard errors for the approximation of the estimates.

**Table 3:** Estimates for segmented linear regression lines for average daily contacts in BC, FHA and VCHA, shown in Figures 1a, 1b and 1c, respectively.

| Parameter | *Regression estimates for average daily contacts* |  |  |
| --- | --- | --- | --- |
|  | BC | FHA | VCHA |
| $\beta_1$ | 0.184** (0.073) | 0.185 (0.126) | 0.111 (0.080) |
| $\beta_2$ | -0.952*** (0.330) | -0.964* (0.568) | -0.745** (0.361) |
| $\beta_3$ | 0.609 (0.506) | 0.766 (0.870) | 0.452 (0.553) |
| $\beta_4$ | 0.111 (0.272) | -0.066 (0.467) | 0.175 (0.297) |
| $\beta_0$ | 5.599*** (0.555) | 5.834*** (0.956) | 5.085*** (0.608) |
| Observations | 40 | 40 | 40 |
| R<sup>2</sup> | 0.643 | 0.384 | 0.482 |
| Adjusted R<sup>2</sup> | 0.603 | 0.313 | 0.423 |
| Residual Std. Error | 0.916 (df = 35) | 1.576 (df = 35) | 1.002 (df = 35) |
| F Statistic | 15.791*** (df = 4; 35) | 5.451*** (df = 4; 35) | 8.149*** (df = 4; 35) |
| *Note:* |  | *p<0.1; **p<0.05; ***p<0.01 |  |

**Table 4:** Estimates for segmented linear regression lines for average daily cases in BC, FHA and VCHA, shown in Figures 1d, 1e and 1f, respectively.

| Parameter | *Regression estimates for average daily cases* |  |  |
| --- | --- | --- | --- |
|  | BC | FHA | VCHA |
| $\beta_1$ | 21.863*** (3.528) | 18.159*** (3.353) | 2.436* (1.244) |
| $\beta_2$ | 103.238*** (15.888) | 63.257*** (15.099) | 32.876*** (5.602) |
| $\beta_3$ | -123.977*** (24.347) | -89.144*** (23.139) | -51.862*** (8.585) |
| $\beta_4$ | -14.387 (13.086) | -4.763 (12.436) | 15.696*** (4.614) |
| $\beta_0$ | 39.411 (26.750) | 1.884 (25.423) | 33.603*** (9.433) |
| Observations | 40 | 40 | 40 |
| R<sup>2</sup> | 0.955 | 0.909 | 0.836 |
| Adjusted R<sup>2</sup> | 0.950 | 0.899 | 0.818 |
| Residual Std. Error | 44.125 (df = 35) | 41.936 (df = 35) | 15.560 (df = 35) |
| F Statistic | 184.430*** (df = 4; 35) | 87.611*** (df = 4; 35) | 44.761*** (df = 4; 35) |
| *Note:* |  | *p<0.1; **p<0.05; ***p<0.01 |  |

**Table 5:** Estimates for segmented linear regression lines for transmission indicator *R*_*t*_ for BC, FHA and VCHA, shown in Figures 1g, 1h and 1i, respectively.

| Parameter | *Regression estimates for transmission indicator $R_t$* |  |  |
| --- | --- | --- | --- |
|  | BC | FHA | VCHA |
| $\beta_1$ | 0.068*** (0.005) | 0.063*** (0.008) | 0.072*** (0.009) |
| $\beta_2$ | -0.139*** (0.024) | -0.168*** (0.036) | -0.097** (0.041) |
| $\beta_3$ | -0.102*** (0.037) | -0.079 (0.055) | -0.174*** (0.063) |
| $\beta_4$ | 0.178*** (0.020) | 0.195*** (0.029) | 0.210*** (0.034) |
| $\beta_0$ | 0.829*** (0.041) | 0.951*** (0.060) | 0.664*** (0.069) |
| Observations | 40 | 40 | 40 |
| $R^2$ | 0.925 | 0.877 | 0.782 |
| Adjusted $R^2$ | 0.916 | 0.862 | 0.757 |
| Residual Std. Error | 0.067 (df = 35) | 0.099 (df = 35) | 0.114 (df = 35) |
| F Statistic | 107.818*** (df = 4;<br>35) | 62.113*** (df = 4;<br>35) | 31.455*** (df = 4;<br>35) |
| *Note:* |  | *p<0.1; **p<0.05; ***p<0.01 |  |

### B Pearson correlation

Correlation analysis is a statistical method for determining the strength of association between two variables. The relationship between the variables is defined by a correlation coefficient *Corr*, which varies between -1 and +1, such that -1 indicates strong negative association while +1 indicates strong positive correlation between the variables [36, 32, 34, 35]. The correlation coefficient of Person correlation of two variables *x* and *z* is given by

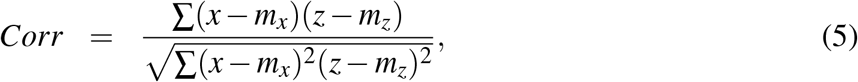

where *m*_*x*_ and *m*_*z*_ are means of *x* and *z*, respectively [35]. In this paper, *x* and *z* represent vectors of time series data with equal lengths. *Corr* is statistically significant if p *< α* = 0.05, otherwise the correlation is insignificant.

### C VAR modeling: stationarization, model fitting, model equations and model estimates

#### C.1 Stationarization of time series for average daily contacts, new cases and *R*_*t*_ in Figure 1

VAR models provide feasible estimates of stationary time series (stationary processes). A stationary process has no time-trends in the mean and variance of the data. Clearly, all time series in Figure 1 are not stationary. Before fitting VAR models, we first stationarize (detrend) the time series by differencing. Differencing works by transforming time series data such that values of the new time series are differences between consecutive values of the original time series. If the resulting time series is not stationary, then differencing can be applied consecutively more than once leading to the second-order difference, third-order difference, etc. We ascertained stationarity of our time series by applying Augmented Dickey-Fuller (ADF) test. According to ADF test, a time series is stationary if p < 0.05. ADF test exists in the *tseries* package in R. Stationary processes of all time series in Figure 1 are presented in Figure 3. Second-order differencing was applied to stationarize time series for average daily contacts, new cases and *R*_*t*_ in BC and VCHA, while third-order differencing was used to stationarize time series for FHA contacts, cases and transmission.

**Figure 3.**
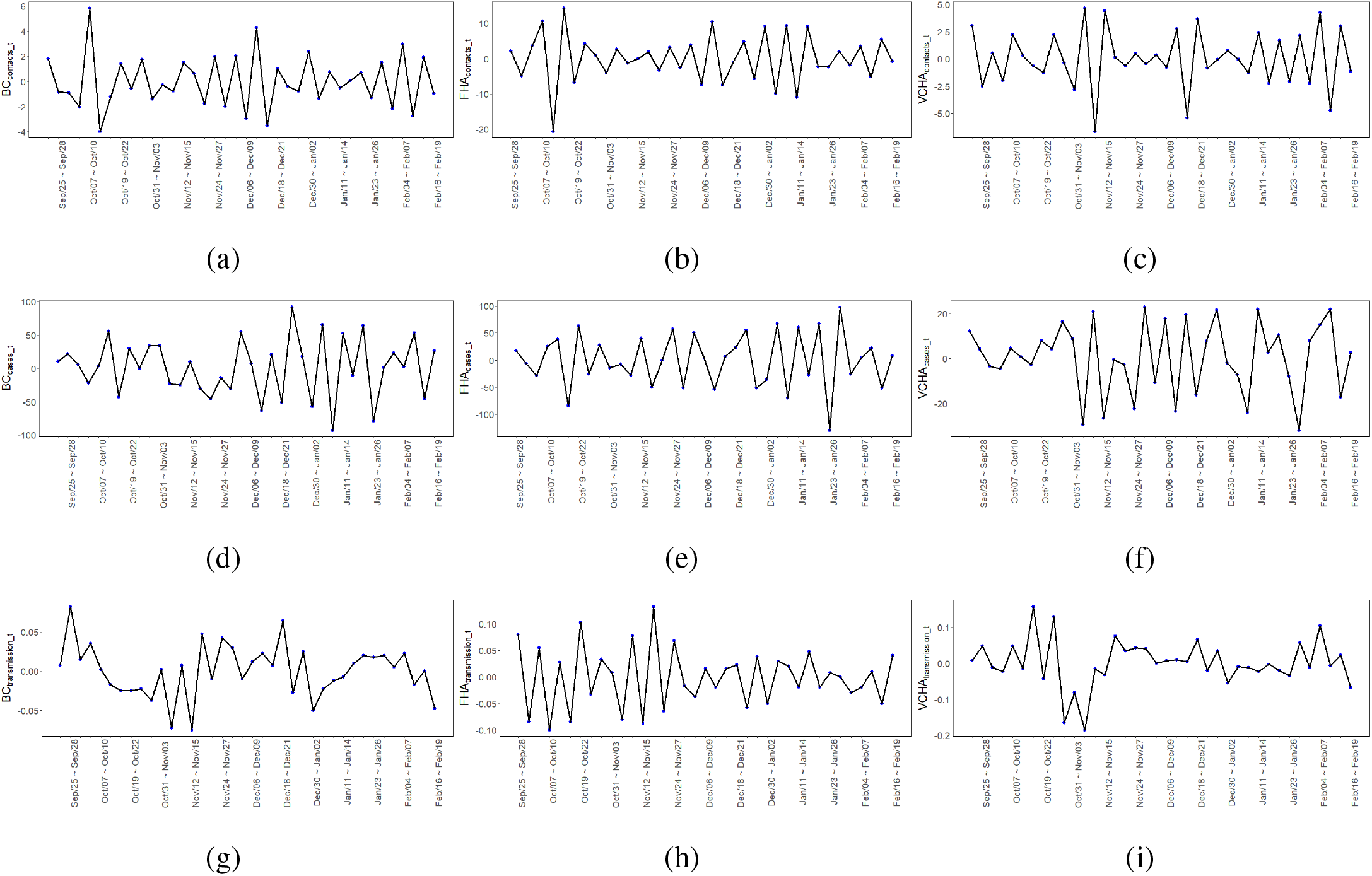
Stationary time series of average daily contacts, average daily cases and *R*_*t*_ in BC (3a, 3d, 3g), FHA (3b, 3e, 3h) and VCHA (3c, 3f, 3i).

#### C.2 VAR model fitting process

We applied the following steps, via the *vars* package in *R*, to fit VAR models of stationary processes of average daily contacts and new cases, and average daily contacts and *R*_*t*_:

i. Use of statistical information criteria (SIC) to determine optimal lag lengths of dependent variables that provide suitable model estimates. The optimal lag lengths are provided by four statistical information criteria, Akaike information criterion (AIC), Hannan Quinn (HQ), Schwarz criterion (SC) and (FPE). In this paper we adopted the lowest suggested lag lengths to derive our VAR model equations.
ii. Estimation of model estimates *µ* and *γ*_*i*_ (see Equation (2)) for each VAR model.
iii. Application of Granger causality test, which is used to determine whether one time series is useful in predicting another. A variable *v* is said to Granger-cause a variable *w* if p < 0.05, otherwise, *v* does not Granger-cause *w*.
iv. Forecast error variance decomposition (FEVD) of model variables. In a FEVD, forecast errors are considered for each equation in the fitted VAR model, then the fitted VAR model is used to determine how much of each error estimate is coming from forecast errors in the other variable. FEVD indicates the amount of information that a variable contributes to step forecast error variance of another variable in the model.

#### C.3 VAR model equations

Here, we derive equations for VAR models for stationary time series of average daily contacts and new cases, and average daily contacts and transmission of COVID-19, in BC, FHA and VCHA, based on the lowest optimal lag lengths suggested by SIC. The lowest suggested lag lengths for our VAR models are presented in Table 6. Therefore, according to Equation (2), the VAR models of *BC*_*contacts*_*t*_ and *BC*_*cases*_*t*_, and *BC*_*contacts*_*t*_ and *BC*_*transmission*_*t*_, are given by

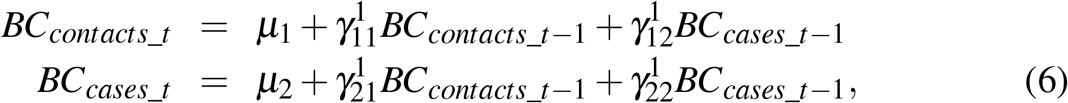

and

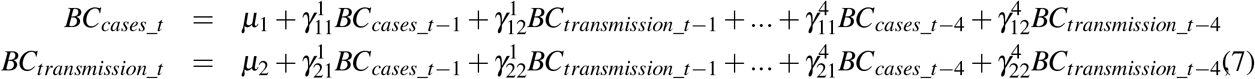

respectively. The VAR models for *FHA*_*contacts*_*t*_ and *FHA*_*cases*_*t*_, *FHA*_*contacts*_*t*_ and *FHA*_*transmission*_*t*_, *VCHA*_*contacts*_*t*_ and *VCHA*_*cases*_*t*_, and *VCHA*_*contacts*_*t*_ and *VCHA*_*transmission*_*t*_ were derived similarly.

**Table 6:** Selected lag lengths of VAR models for stationary time series of average daily contacts and new cases, and average daily contacts and new cases in BC, FHA and VCHA.

| VAR model variables | Lag length | SIC |
| --- | --- | --- |
| $BC_{contacts\_t}$ vs. $BC_{cases\_t}$ | 1 | AIC, HQ, SC, FPE |
| $BC_{contacts\_t}$ vs. $BC_{transmission\_t}$ | 4 | SC |
| $FHA_{contacts\_t}$ vs. $FHA_{cases\_t}$ | 6 | FPE |
| $FHA_{contacts\_t}$ vs. $FHA_{transmission\_t}$ | 9 | FPE |
| $VCHA_{contacts\_t}$ vs. $VCHA_{cases\_t}$ | 2 | SC |
| $VCHA_{contacts\_t}$ vs. $VCHA_{transmission\_t}$ | 10 | AIC, HQ, SC, FPE |

#### C.4 Estimates for VAR models of average daily contacts, new cases and *R*_*t*_

**Table 7:** Estimation results for VAR(1) model of variables *BC*_*contacts*_*t*_ and *BC*_*cases*_*t*_

|  | *Dependent variable:* |  |
| --- | --- | --- |
| | $BC_{contacts\_t}$ | $BC_{cases\_t}$ |
| $BC_{contacts\_t-1}$ | -0.770***<br>(0.112) | 7.930***<br>(2.804) |
| $BC_{cases\_t-1}$ | -0.011*<br>(0.005) | -0.400***<br>(0.137) |
| Observations | 37 | 37 |
| R<sup>2</sup> | 0.577 | 0.375 |
| Adjusted R<sup>2</sup> | 0.553 | 0.339 |
| Residual Std. Error (df = 35) | 1.399 | 35.071 |
| F Statistic (df = 2; 35) | 23.850*** | 10.504*** |

*Note:* \*p<0.1; \*\*p<0.05; \*\*\*p<0.01

**Table 8:** Estimation results for VAR(4) model of variables *BC*_*contacts*_*t*_ and *BC*_*tranmission*_*t*_

|  | *Dependent variable:* |  |
| --- | --- | --- |
| | $BC_{contacts\_t}$ | $BC_{tranmission\_t}$ |
| $BC_{contacts\_t-1}$ | -1.482***<br>(0.144) | 0.001<br>(0.004) |
| $BC_{tranmission\_t-1}$ | 7.907<br>(6.886) | -0.018<br>(0.171) |
| $BC_{contacts\_t-2}$ | -1.283***<br>(0.215) | 0.003<br>(0.005) |
| $BC_{tranmission\_t-2}$ | -5.839<br>(6.836) | 0.832***<br>(0.170) |
| $BC_{contacts\_t-3}$ | -1.008***<br>(0.213) | 0.002<br>(0.005) |
| $BC_{tranmission\_t-3}$ | -7.574<br>(6.338) | -0.116<br>(0.158) |
| $BC_{contacts\_t-4}$ | -0.493***<br>(0.146) | 0.002<br>(0.004) |
| $BC_{tranmission\_t-4}$ | 8.520<br>(6.429) | -0.463***<br>(0.160) |
| Observations | 34 | 34 |
| R<sup>2</sup> | 0.837 | 0.528 |
| Adjusted R<sup>2</sup> | 0.786 | 0.383 |
| Residual Std. Error (df = 26) | 0.990 | 0.025 |
| F Statistic (df = 8; 26) | 16.643*** | 3.639*** |
| *Note:* | *p<0.1; **p<0.05; ***p<0.01 |  |

**Table 9:** Estimation results for VAR(6) model of variables *FHA*_*contacts*_*t*_ and *FHA*_*cases*_*t*_

|  | *Dependent variable:* |  |
| --- | --- | --- |
| | $FHA_{contacts\_t}$ | $FHA_{cases\_t}$ |
| $FHA_{contacts\_t-1}$ | −1.937***<br>(0.152) | −1.134<br>(3.630) |
| $FHA_{cases\_t-1}$ | 0.004<br>(0.009) | −1.182***<br>(0.223) |
| $FHA_{contacts\_t-2}$ | −2.051***<br>(0.289) | −5.295<br>(6.906) |
| $FHA_{cases\_t-2}$ | 0.024*<br>(0.014) | −1.355***<br>(0.328) |
| $FHA_{contacts\_t-3}$ | −1.886***<br>(0.332) | −5.200<br>(7.931) |
| $FHA_{cases\_t-3}$ | 0.021<br>(0.017) | −1.128**<br>(0.415) |
| $FHA_{contacts\_t-4}$ | −1.497***<br>(0.306) | −6.283<br>(7.301) |
| $FHA_{cases\_t-4}$ | 0.022<br>(0.017) | −0.945**<br>(0.406) |
| $FHA_{contacts\_t-5}$ | −0.925***<br>(0.220) | −4.017<br>(5.254) |
| $FHA_{cases\_t-5}$ | 0.018<br>(0.015) | −0.608<br>(0.367) |
| $FHA_{contacts\_t-6}$ | −0.407***<br>(0.107) | −2.137<br>(2.559) |
| $FHA_{cases\_t-6}$ | −0.017<br>(0.010) | −0.173<br>(0.249) |
| Observations | 31 | 31 |
| R<sup>2</sup> | 0.969 | 0.779 |
| Adjusted R<sup>2</sup> | 0.949 | 0.639 |
| Residual Std. Error (df = 19) | 1.267 | 30.241 |
| F Statistic (df = 12; 19) | 48.749*** | 5.571*** |
| *Note:* |  |  |
| *p<0.1; **p<0.05; ***p<0.01 |  |  |

**Table 10:**
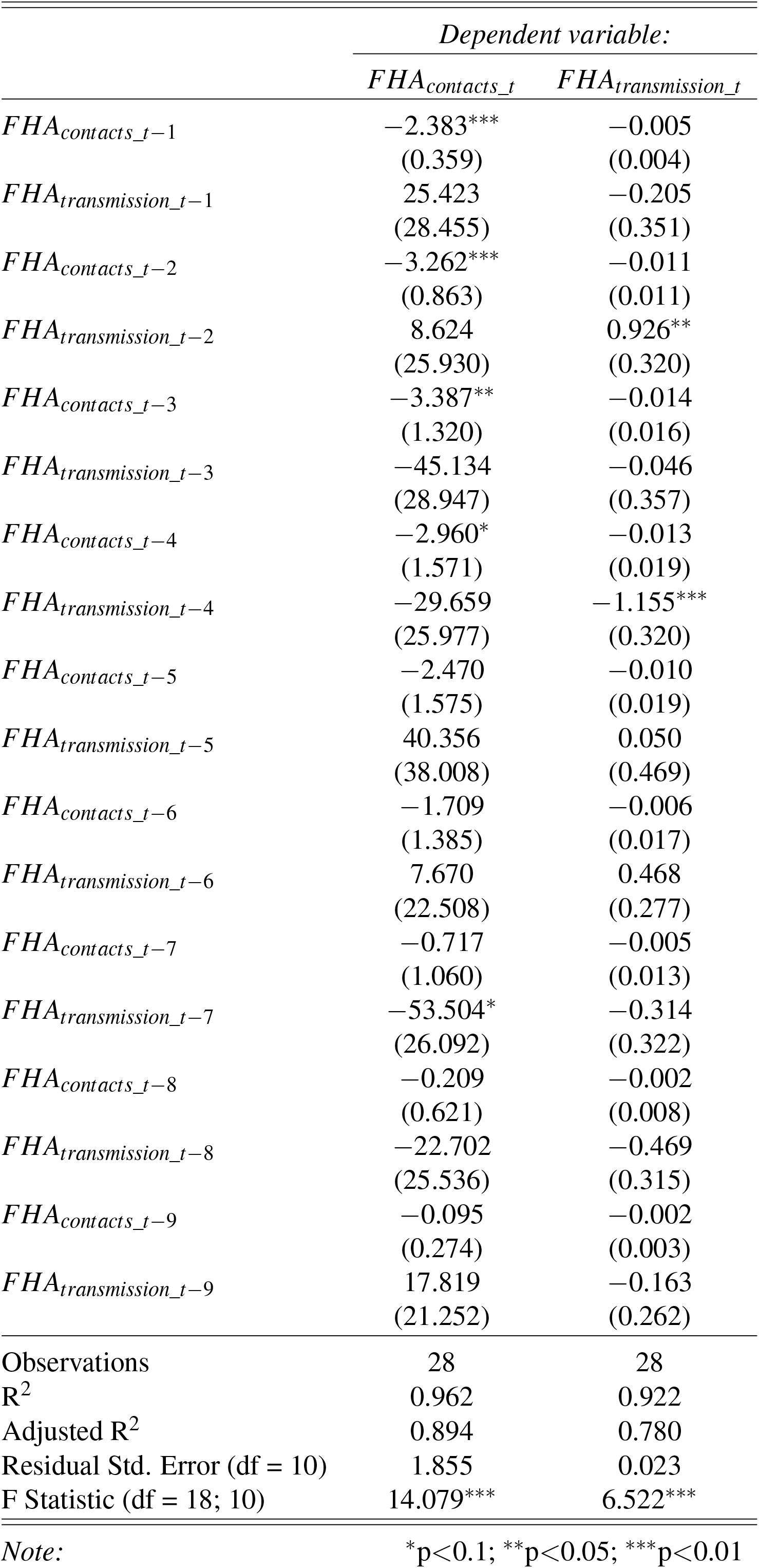
Estimation results for VAR(9) model of variables *FHA*_*contacts*_*t*_ and *FHA*_*transmission*_*t*_

|  | *Dependent variable:* |  |
| --- | --- | --- |
| | $FHA_{contacts\_t}$ | $FHA_{transmission\_t}$ |
| $FHA_{contacts\_t-1}$ | -2.383***<br>(0.359) | -0.005<br>(0.004) |
| $FHA_{transmission\_t-1}$ | 25.423<br>(28.455) | -0.205<br>(0.351) |
| $FHA_{contacts\_t-2}$ | -3.262***<br>(0.863) | -0.011<br>(0.011) |
| $FHA_{transmission\_t-2}$ | 8.624<br>(25.930) | 0.926**<br>(0.320) |
| $FHA_{contacts\_t-3}$ | -3.387**<br>(1.320) | -0.014<br>(0.016) |
| $FHA_{transmission\_t-3}$ | -45.134<br>(28.947) | -0.046<br>(0.357) |
| $FHA_{contacts\_t-4}$ | -2.960*<br>(1.571) | -0.013<br>(0.019) |
| $FHA_{transmission\_t-4}$ | -29.659<br>(25.977) | -1.155***<br>(0.320) |
| $FHA_{contacts\_t-5}$ | -2.470<br>(1.575) | -0.010<br>(0.019) |
| $FHA_{transmission\_t-5}$ | 40.356<br>(38.008) | 0.050<br>(0.469) |
| $FHA_{contacts\_t-6}$ | -1.709<br>(1.385) | -0.006<br>(0.017) |
| $FHA_{transmission\_t-6}$ | 7.670<br>(22.508) | 0.468<br>(0.277) |
| $FHA_{contacts\_t-7}$ | -0.717<br>(1.060) | -0.005<br>(0.013) |
| $FHA_{transmission\_t-7}$ | -53.504*<br>(26.092) | -0.314<br>(0.322) |
| $FHA_{contacts\_t-8}$ | -0.209<br>(0.621) | -0.002<br>(0.008) |
| $FHA_{transmission\_t-8}$ | -22.702<br>(25.536) | -0.469<br>(0.315) |
| $FHA_{contacts\_t-9}$ | -0.095<br>(0.274) | -0.002<br>(0.003) |
| $FHA_{transmission\_t-9}$ | 17.819<br>(21.252) | -0.163<br>(0.262) |
| Observations | 28 | 28 |
| R<sup>2</sup> | 0.962 | 0.922 |
| Adjusted R<sup>2</sup> | 0.894 | 0.780 |
| Residual Std. Error (df = 10) | 1.855 | 0.023 |
| F Statistic (df = 18; 10) | 14.079*** | 6.522*** |

Note:
\*p<0.1; \*\*p<0.05; \*\*\*p<0.01

**Table 11:**
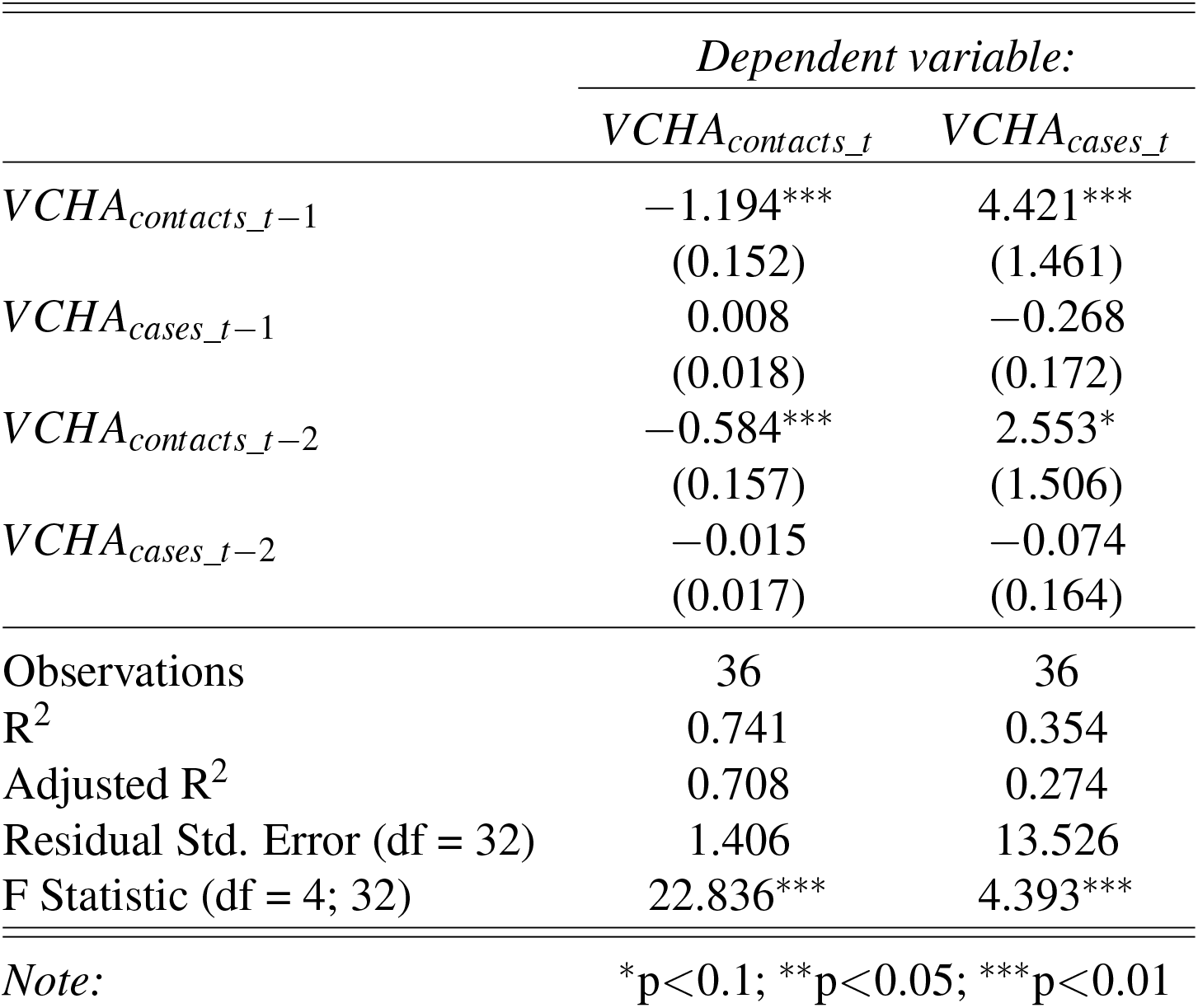
Estimation results for VAR(2) model of variables *VCHA*_*contacts*_*t*_ and *VCHA*_*cases*_*t*_

**Table 12:** Estimation results for VAR(10) model of variables *VCHA*_*contacts*_*t*_ and *VCHA*_*transmission*_*t*_

|  | *Dependent variable:* |  |
| --- | --- | --- |
| | $VCHA_{contacts\_t}$ | $VCHA_{transmission\_t}$ |
| $VCHA_{contacts\_t-1}$ | -1.395***<br>(0.274) | 0.010<br>(0.010) |
| $VCHA_{transmission\_t-1}$ | 5.984<br>(7.293) | 0.284<br>(0.258) |
| $VCHA_{contacts\_t-2}$ | -1.090*<br>(0.507) | 0.007<br>(0.018) |
| $VCHA_{transmission\_t-2}$ | -5.706<br>(6.215) | 0.589**<br>(0.220) |
| $VCHA_{contacts\_t-3}$ | -0.706<br>(0.565) | -0.007<br>(0.020) |
| $VCHA_{transmission\_t-3}$ | 10.938<br>(8.920) | -0.508<br>(0.316) |
| $VCHA_{contacts\_t-4}$ | -0.643<br>(0.555) | -0.002<br>(0.020) |
| $VCHA_{transmission\_t-4}$ | -0.152<br>(9.099) | -0.673*<br>(0.322) |
| $VCHA_{contacts\_t-5}$ | -0.693<br>(0.577) | -0.007<br>(0.020) |
| $VCHA_{transmission\_t-5}$ | -12.496<br>(10.769) | 0.503<br>(0.381) |
| $VCHA_{contacts\_t-6}$ | -0.244<br>(0.601) | 0.004<br>(0.021) |
| $VCHA_{transmission\_t-6}$ | -1.131<br>(10.323) | 0.186<br>(0.366) |
| $VCHA_{contacts\_t-7}$ | -0.560<br>(0.542) | 0.015<br>(0.019) |
| $VCHA_{transmission\_t-7}$ | 14.575<br>(9.494) | -0.664*<br>(0.336) |
| $VCHA_{contacts\_t-8}$ | -0.511<br>(0.544) | 0.026<br>(0.019) |
| $VCHA_{transmission\_t-8}$ | -1.761<br>(9.496) | 0.167<br>(0.336) |
| $VCHA_{contacts\_t-9}$ | -0.200<br>(0.472) | 0.020<br>(0.017) |
| $VCHA_{transmission\_t-9}$ | -15.999**<br>(5.955) | -0.127<br>(0.211) |
| $VCHA_{contacts\_t-10}$ | 0.085<br>(0.240) | 0.005<br>(0.009) |
| $VCHA_{transmission\_t-10}$ | 13.138*<br>(6.293) | -0.137<br>(0.223) |
| Observations | 28 | 28 |
| R<sup>2</sup> | 0.982 | 0.938 |
| Adjusted R<sup>2</sup> | 0.936 | 0.781 |
| Residual Std. Error (df = 8) | 0.721 | 0.026 |
| F Statistic (df = 20; 8) | 21.482*** | 6.006*** |

Note:
\*p<0.1; \*\*p<0.05; \*\*\*p<0.01

